# Outcomes of Fibrate vs Statin Therapy in Patients With Nonproliferative Diabetic Retinopathy and Type 2 Diabetes

**DOI:** 10.1101/2025.08.12.25333543

**Authors:** Luke Nelson, Jesse Maynard, Matthew Morckos, Rubens Petit Homme, Samrawit Zinabu, Rawan Elkomi, Miriam Michael, Salman Yousuf

**Affiliations:** Howard University College of Medicine, Department of Ophthalmology, Washington, D.C

**Author notes:** Corresponding Author: Luke Nelson, 2041 Georgia Ave NW, Tower 2nd Floor, Suite 2100, Washington, DC 20060.

## Abstract

**Importance:** Diabetic retinopathy (DR) progression significantly impacts vision and quality of life in patients with Type 2 Diabetes Mellitus (T2DM). Statins and fibrates are commonly prescribed lipid-lowering medications, but their comparative effectiveness in DR progression remains uncertain.

**Objective:** To determine whether fibrates reduce the risk of DR progression compared to statins in patients with T2DM and nonproliferative diabetic retinopathy (NPDR).

**Design, Setting, and Participants:** Retrospective cohort study using the TriNetX Global Collaborative Network, a multicentered, population-based electronic medical record database.

Inclusion criteria consisted of patients diagnosed with T2DM and NPDR 5-20 years prior. Patients were propensity score matched based on demographics and comorbidities. Two cohorts were defined: patients on fibrates but not statins (n = 543), and patients on statins but not fibrates (n = 60,135). After matching, each cohort included 542 patients.

**Main Outcomes and Measures:** Primary outcomes: intravitreal antiVEGF injection or retinal laser procedures

Secondary outcomes: progression to PDR, vitreous hemorrhage (VH), neovascularization (NV), or tractional retinal detachment (TRD)

Tertiary outcomes: neovascular glaucoma (NVG) or pars plana vitrectomy (PPV).

**Results:** Fibrate-treated patients had a 57.3% risk reduction in anti-VEGF injection (RR = 0.427; 95% CI: 0.219, 0.830; p < 0.011) and longer time-to-injection (log-rank x^2^ = 4.927; p < 0.027). PDR risk was reduced by 59.3% (RR = 0.407; 95% CI: 0.242, 0.684; p < 0.001) with delayed progression (log-rank x^2^ = 8.657; p < 0.004).

Fibrate use was associated with 72.1% lower instantaneous risk of NGV (HR = 0.279; 95% CI: 0.079, 0.991, p < 0.015) and delayed onset (log-rank x^2^ = 4.448, p < 0.036) despite a higher survival probability in the statin group (97.08% vs 96.65%).

Fibrates lowered the absolute risk of NV (−0.019; 95% CI: −0.030, −0.007; p < 0.002) but increased risk of TRD (0.018; 95% CI: 0.007, 0.030; p < 0.003); however, neither occurred in the comparison group, limiting statistical power.

**Conclusions and Relevance:** Among patients with T2DM and NPDR, fibrates were associated with reduced risk of antiVEGF injection, PDR progression, and NVG onset compared to statins. These findings suggest fibrates may help mitigate DR progression.

## Introduction

Diabetic retinopathy (DR) is a common microvascular complication of diabetes mellitus and a leading cause of vision loss in working-age adults.^1^ Chronic hyperglycemia drives DR through mechanisms like retinal microvascular damage, inflammation, and ischemia-induced growth factor release (e.g. VEGF), leading to features from non-proliferative DR (microaneurysms, hemorrhages, hard exudates) to sight-threatening proliferative DR and macular edema.^1^ While tight glycemic and blood pressure control remain the cornerstone of DR prevention, dyslipidemia has been implicated as an additional risk factor for DR progression, though findings have been somewhat conflicting.^2^ Hard exudates in the retina (lipid-rich deposits) suggest that abnormal lipid profiles might exacerbate retinal vascular leakage and edema. This possibility has prompted interest in lipid-lowering therapies – notably statins and fibrates – as potential modulators of DR progression beyond their cardiovascular benefits.

Statins 3-Hydroxy-3-Methylglutaryl-Coenzyme A (HMG-CoA) reductase inhibitors and fibrates, Peroxisome proliferator-activated receptor alpha (PPAR-α) agonists like fenofibrate, are widely used to manage dyslipidemia in patients with type 2 diabetes. Both classes have pleiotropic effects that could influence the retinal microvasculature. Over the past two decades, numerous studies – including observational analyses, randomized trials, and animal experiments – have examined whether these agents can slow DR progression or prevent vision-threatening complications. Notably, two large clinical trials (FIELD and ACCORD Eye) provided seminal evidence that fenofibrate slows DR progression, whereas evidence for statins has been less clear.^1^ In this report, we provide a structured review of the comparative impact of statins versus fibrates on diabetic retinopathy. We discuss clinical outcomes from trials (statins vs fibrates, fibrates vs placebo, statins vs placebo), summarize observational and preclinical findings on DR outcomes (e.g. progression to proliferative DR, need for laser photocoagulation or intravitreal therapy), and explore the molecular mechanisms that might explain differences between these drug classes.

In this report, we leveraged a large, international database to directly compare the effect of statins versus fibrates on the progression of diabetic retinopathy in real-world clinical settings. By integrating a literature review with original data analysis, this study provides novel insights into how these widely used lipid-lowering therapies may differentially influence DR progression. Potential molecular mechanisms that might explain differences between these drug classes are also explored.

## Methods

### Study Design and Population

This retrospective, comparative cohort study used the TriNetX Global Collaborative Network utilizing de-identified electronic medical records from 149 healthcare organizations (HCOs) across 21 countries. Data use was determined to be exempt from institutional review board oversight by both Western IRB and MetroHealth, in accordance with Section §164.514(b)(1) of the HIPAA Privacy Rule, as reviewed by a qualified expert. This study adhered to the Strengthening the Reporting of Observational Studies in Epidemiology (STROBE) guidelines.

The analysis was conducted on July 1, 2025, and included adults with type 2 diabetes mellitus (T2DM) and any stage of nonproliferative diabetic retinopathy (NPDR) with diagnosis occurring at least 5 years prior to cohort entry. The index event for inclusion was defined as the first recorded diagnosis of any stage of NPDR with or without DME and were excluded if they had any prior diagnosis of: type 1 diabetes, exudative age-related macular degeneration (wet AMD), serous retinal detachment, separation of retinal layers, retinal detachment and breaks, cystoid macular degeneration, solar retinopathy, hypertensive retinopathy, other non-diabetic proliferative retinopathy, retinal vascular occlusions, sickle-cell disorders, degenerative myopia with choroidal neovascularization. Patients with index events occurring more than 20 years prior were also excluded.

Two cohorts were defined: patients taking statins without fibrates (n = 60,135), and patients taking fibrates without statins (n = 543). Medication use was based on any documented prescription on or after index date. To isolate the ophthalmic effects of each lipid-lower drug (LLD) class, patients with concurrent post-index use of both statins and fibrates were excluded.

To minimize potential confounding factors, patients were 1:1 propensity score matched and balanced on the following characteristics: overall age, age at index event, gender, race, and ethnicity as shown in Table 1. Patients were also matched on relative serum cholesterol (triglycerides, LDL, HDL) and HbA1c. Standardized differences < 0.1 post-matching were considered acceptable, as shown in Figure 1, emphasizing the distribution of propensity scores before and after matching.

**Figure 1:**
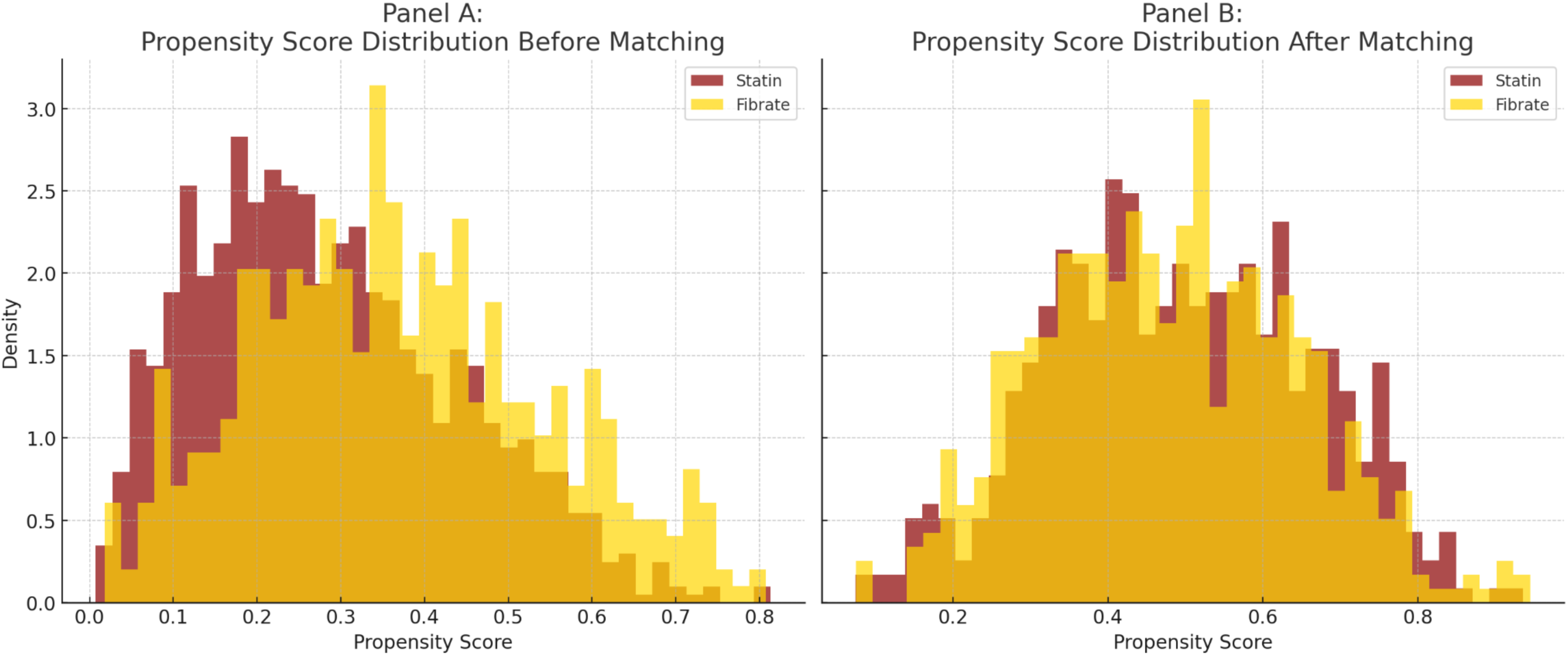
Distribution of Propensity Scores Before and After Matching. Panel A shows the density distribution of propensity scores prior to matching, highlighting baseline differences between statin (burgundy) and fibrate (gold) cohorts. Panel B shows the post-matching distributions, demonstrating improved balance across cohorts.

**Table 1:** Baseline Characteristics of Patients Before and After Propensity Score Matching. Demographic characteristics of patients with type 2 diabetes and nonproliferative diabetic retinopathy treated with statins or fibrates, before and after 1:1 propensity score matching (n = 542 per group). Before matching, groups differed in age, sex, and race. After matching, no significant differences remained, and standardized differences were minimized, indicating successful covariate balance.

| Characteristic | Before Matching<br>(% Cohort) |  |  |  | After<br>Matching<br>(% Cohort) |  |  |  |
| --- | --- | --- | --- | --- | --- | --- | --- | --- |
|  | Statin<br>(n = 60,135) | Fibrate<br>(n = 543) | P-Value | Std<br>Diff | Statin<br>(n = 542) | Fibrate<br>(n = 542) | P-Value | Std<br>Diff |
| **Demographics** |  |  |  |  |  |  |  |  |
| Age at index ± mean<br>(SD) | 64.9 ± 11.4<br>(100%) | 62.7 ± 13.4<br>(100%) | <0.001 | 0.146 | 63.6 ± 12.3<br>(100%) | 62.7 ± 13.4<br>(100%) | 0.280 | 0.066 |
| Female, n (%) | 29,985 (49.9%) | 238 (43.8%) | 0.005 | 0.121 | 242 (44.6%) | 237 (43.7%) | 0.760 | 0.019 |
| Male, n (%) | 29,177 (48.5%) | 291 (53.6%) | 0.019 | 0.102 | 285 (52.6%) | 291 (53.7%) | 0.715 | 0.022 |
| White, n (%) | 32,402 (53.9%) | 358 (65.9%) | <0.001 | 0.248 | 360 (66.4%) | 357 (65.9%) | 0.847 | 0.012 |
| Black or African<br>American, n (%) | 13,722 (22.8%) | 39 (7.2%) | <0.001 | 0.449 | 34 (6.3%) | 39 (7.2%) | 0.545 | 0.037 |
| Asian, n (%) | 4,397 (7.3%) | 53 (9.8%) | 0.029 | 0.088 | 56 (10.3%) | 53 (9.8%) | 0.762 | 0.018 |
| Hispanic or Latino | 9,257 (15.4%) | 94 (17.3%) | 0.218 | 0.052 | 86 (15.9%) | 94 (17.3%) | 0.514 | 0.040 |
| Not Hispanic or<br>Latino | 40,214 (66.9%) | 311 (57.3%) | <0.001 | 0.199 | 312 (57.6%) | 311 (57.4%) | 0.951 | 0.004 |
| Native Hawaiian or<br>Other Pacific Islander | 1,189 (2.0%) | 10 (1.8%) | 0.821 | 0.010 | 10 (1.8%) | 10 (1.8%) | 1 | <0.001 |
| American Indian or<br>Alaska Native | 315 (0.5%) | 10 (1.8%) | <0.001 | 0.122 | 10 (1.8%) | 10 (1.8%) | 1 | <0.001 |

### Outcomes

Primary outcomes included intravitreal antiVEGF injection, panretinal photocoagulation (PRP), or macular laser procedures. Secondary outcomes included diagnosis of proliferative diabetic retinopathy (PDR), vitreous hemorrhage (VH), neovascularization (NV), and tractional retinal detachment (TRD). Tertiary outcomes included pars plana vitrectomy (PPV) and progression to neovascular glaucoma (NVG).

Potential adverse outcomes were also measured and included serum creatinine, aspartate aminotransferase (AST), alanine aminotransferase (ALT), and occurrences of acute kidney injury (AKI). All outcomes were identified using diagnosis and procedure codes captured in the post-index window, which began on index date.

Patients were excluded from the outcome analysis if they experienced a given outcome prior to the index date.

### Statistical Analysis

Differences in categorical variables were compared using risk differences and risk ratio (RR) with 95% confidence intervals (CI), and were evaluated via a two-tailed z-test.

Kaplan-Meier survival curves were used to estimate time to diagnosis or intervention, and were evaluated using the standard log-rank test. Hazard ratios (HR) assessed instantaneous risk at any timepoint and were calculated using Cox proportional hazards regression.

All analyses were conducted using TriNetX’s embedded analytics tools and any p-value < 0.05 was considered statistically significant.

## Results

### Proliferative Diabetic Retinopathy

Patients in the fibrate group had a 59.3% risk reduction in PDR diagnosis compared to the statin group (RR = 0.407; 95% CI: 0.242, 0.684; p < 0.001) as shown in Figure 2. Survival was also increased (log-rank x^2^ = 8.657; p < 0.004) but without any changes in HR.

**Figure 2:**
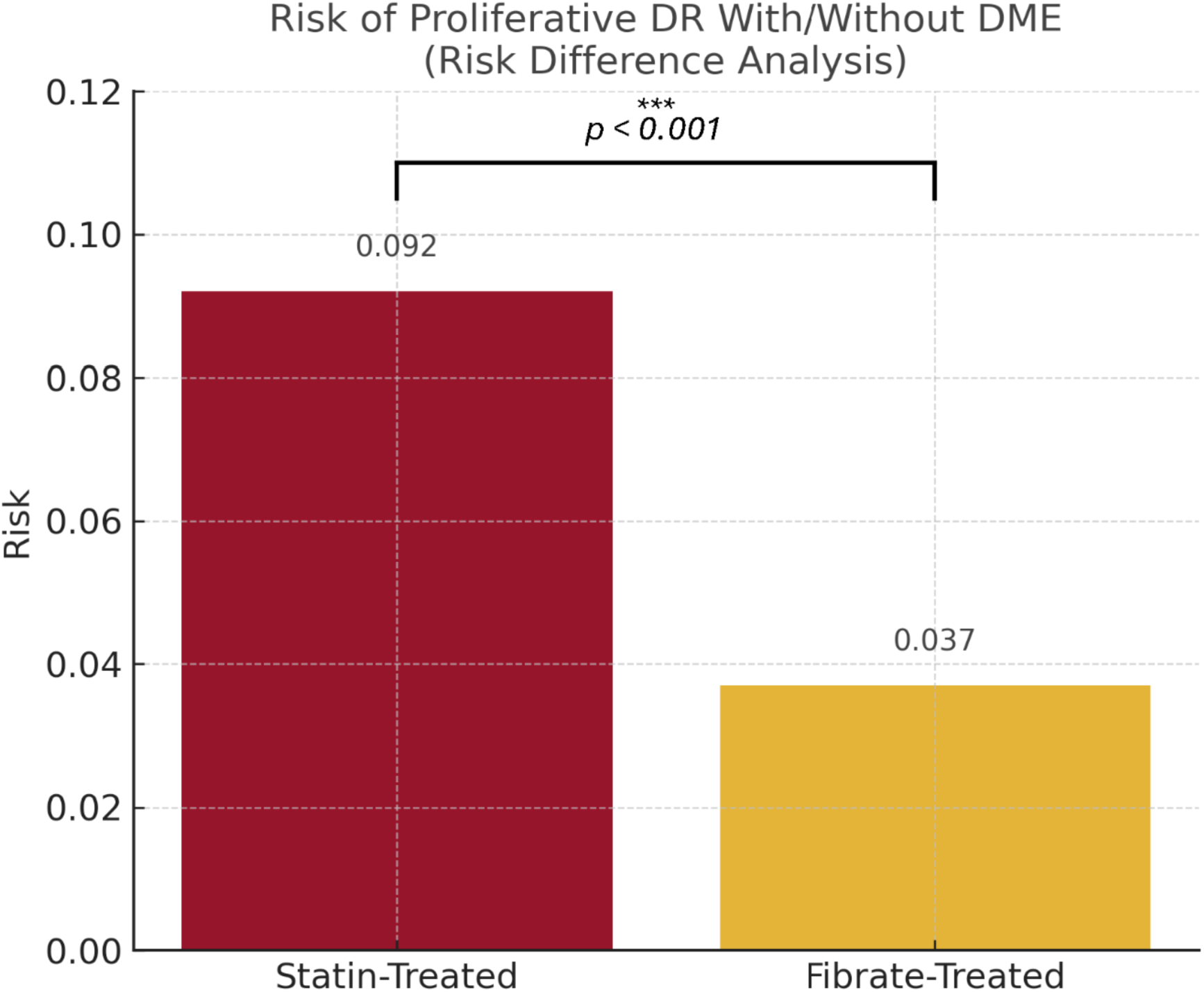
Risk of Proliferative Diabetic Retinopathy (PDR) With or Without Diabetic Macular Edema (DME) Among Statin- vs Fibrate-Treated Patients. This bar graph illustrates the absolute risk of developing PDR (with or without DME) among patients with type 2 diabetes and nonproliferative diabetic retinopathy treated with statin or fibrate monotherapy. Patients treated with statins had a significantly higher risk (9.2%) compared to those on fibrates (3.7%), corresponding to a risk difference of −5.5% (95% CI, −8.5% to −2.4%; *p* < 0.001). Bars represent absolute risk; horizontal line denotes statistically significant difference in proportions.

### Intravitreal AntiVEGF Injection

Fibrate-treated patients had a 57.3% risk reduction of anti-VEGF injection (RR = 0.427; 95% CI: 0.219, 0.830; p < 0.011) and higher survival (log-rank x^2^ = 4.927; p < 0.027) with no change in HR (Figure 3).

**Figure 3:**
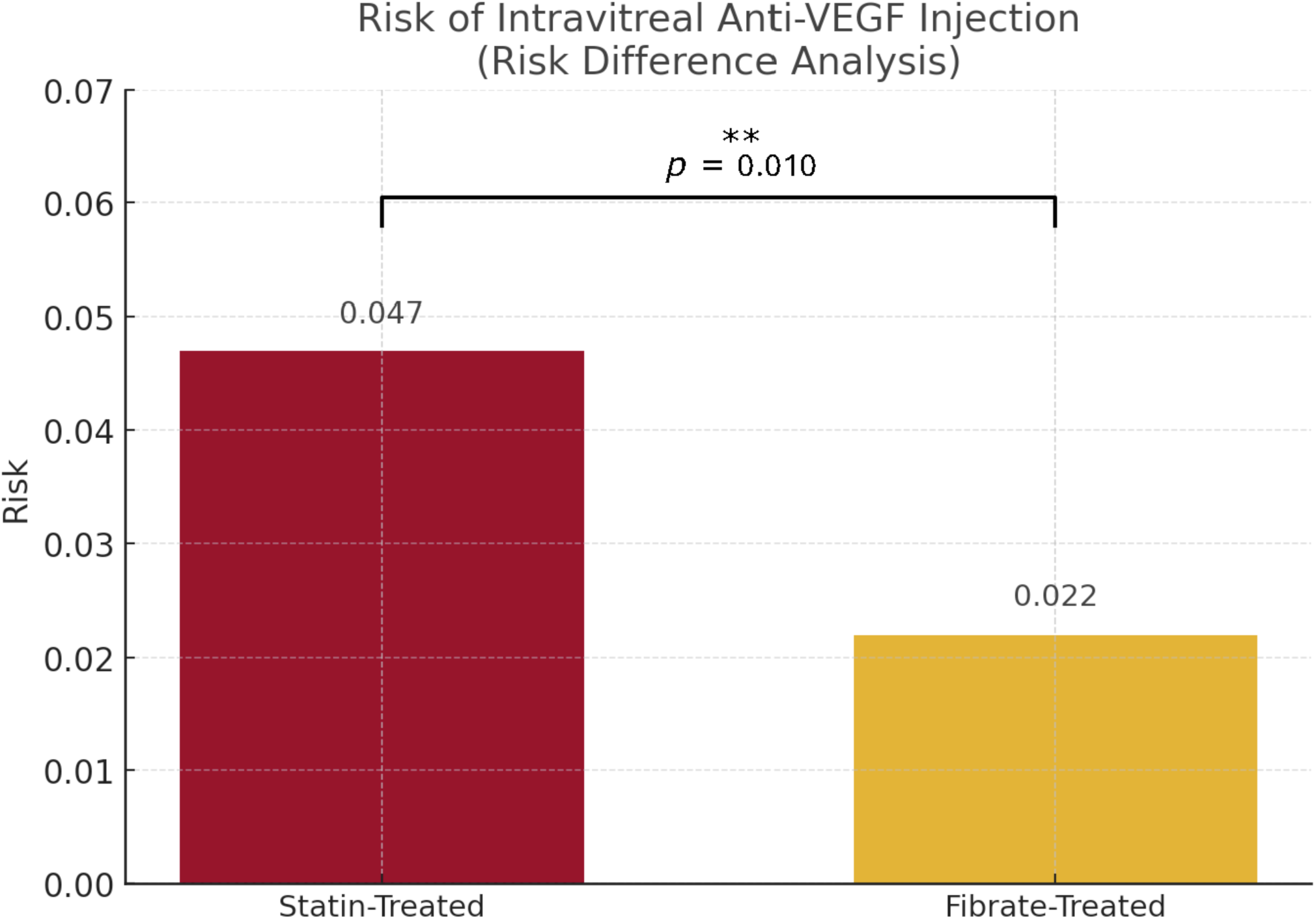
Risk of Intravitreal Anti-VEGF Injection Among Statin- and Fibrate-Treated Patients. This bar plot displays the absolute risk of requiring intravitreal anti–vascular endothelial growth factor (VEGF) injections among patients with type 2 diabetes and nonproliferative diabetic retinopathy (NPDR), stratified by lipid-lowering therapy. The risk was significantly higher in the statin-treated group (4.7%) compared to the fibrate-treated group (2.2%), resulting in an absolute risk difference of −2.5% (95% CI, −4.8% to −0.2%; *p* = 0.036). These findings suggest a potential protective role of fibrates against vision-threatening disease progression necessitating anti-VEGF therapy.

### Neovascularization, Tractional Retinal Detachment, and Neovascular Glaucoma

Fibrate users demonstrated a reduced absolute risk of NV (risk difference = −0.019; 95% CI: −0.030, −0.007; p < 0.002) but an increased absolute risk of TRD (risk difference = 0.018; 95% CI: 0.007, 0.030; p < 0.003) as shown in Figure 4. However, neither outcome occurred in the comparison group, limiting statistical power and preventing the calculation of RRs, Kaplan-Meier estimates, and HRs.

**Figure 4:**
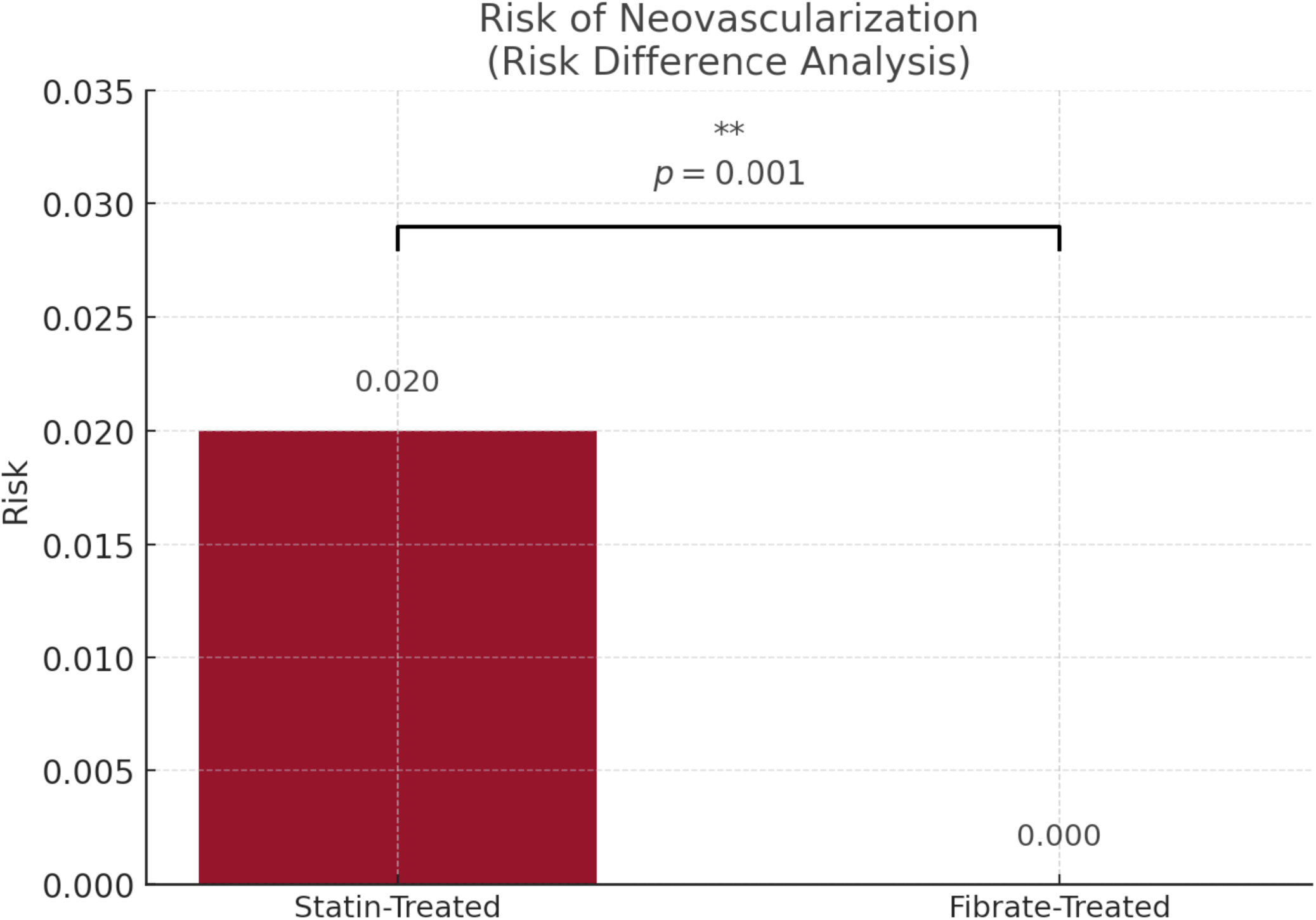
Risk of Neovascularization in Mild NPDR by Treatment Type. The proportion of patients developing retinal neovascularization after exclusion of those with prior outcomes is shown. Statin-treated patients had a 2.0% risk, whereas no events were observed in the fibrate group. The resulting risk difference was –2.0% (95% CI: – 3.2% to –0.8%, p = 0.001), ** indicating a statistically significant reduction with fibrate therapy.

Patients taking fibrates also demonstrated a 72.1% reduction in instantaneous risk for NVG (HR = 0.279; 95% CI: 0.079, 0.991, p < 0.015) and enhanced time-to-event (log-rank x^2^ = 4.448, p < 0.036), despite similar overall incidence and slightly higher absolute survival probability in the statin group (97.08% vs 96.65%).

### Panretinal Photocoagulation, Macular Laser, Vitreous Hemorrhage, and Pars Plana Vitrectomy

No significant differences were observed between groups for PRP, VH, or PPV. For macular laser treatment, Statin monotherapy was associated with a significantly increased hazard of undergoing macular laser treatment (HR = 1.42; 95% CI, 0.32–6.36; p = 0.024), as shown in forest plot in Figure 5.

**Figure 5:**
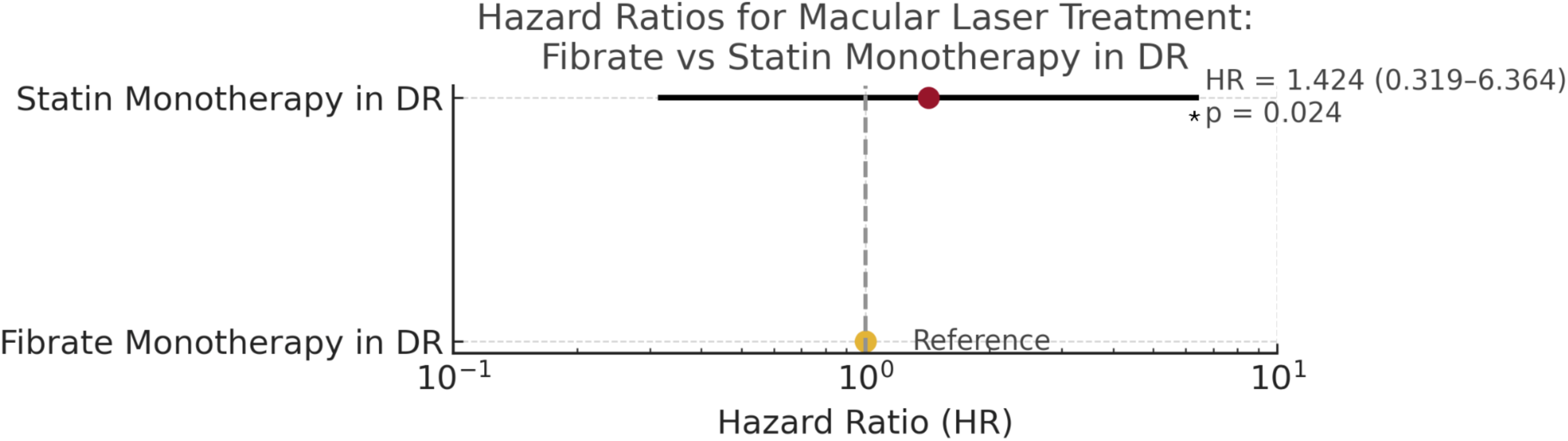
Forest plot comparing hazard ratios for macular laser treatment in patients with diabetic retinopathy (DR) treated with statin versus fibrate monotherapy. Hazard ratios (HRs) and 95% confidence intervals (CIs) are shown on a logarithmic scale. Fibrate monotherapy served as the reference group (HR = 1.0). Statin monotherapy was associated with a significantly increased hazard of undergoing macular laser treatment (HR = 1.42; 95% CI, 0.32–6.36; p = 0.024). The dashed vertical line at HR = 1.0 represents the null value

### Adverse Outcomes: Creatinine, ALT, AST, AKI

Statin users had a 28.2% greater risk for AKI diagnosis (RR = 0.718; 95% CI: 0.551, 0.936; p < 0.014) but no difference in survival or HR. No significant differences were observed in creatinine, AST, or ALT levels.

## Discussion

This study found that fibrate use in patients with NPDR was associated with a significantly lower risk of progression to proliferative diabetic retinopathy, fewer instances of intravitreal anti-VEGF injections, and delayed onset of NVG diagnosis. Fibrates also reduced the absolute risk of NV but were linked to higher absolute risk of TRD, despite no event occurrence in the comparison group.

The findings above highlight a valuable opportunity to repurpose cardiovascular drugs for ocular benefit in diabetes.^1^ Fenofibrate’s consistent benefit on diabetic retinopathy – demonstrated in randomized trials and corroborated by real-world evidence – represents one of the most significant advances in the medical (non-surgical) management of DR in recent decades. It underscores that DR progression is not driven by hyperglycemia alone, but also by dyslipidemia and inflammation that can be targeted. The fact that fenofibrate’s effect was independent of blood sugar suggests that even diabetics with optimally controlled blood sugar might further reduce their DR risk with this therapy.^3^ Consequently, some clinical guidelines have endorsed fenofibrate for DR management. For example, Australia approved fenofibrate for T2D patients with DR to slow progression,^4^ and the UK’s NICE guidelines note fenofibrate’s benefit in people with type 2 diabetes who have existing retinopathy.^5^ This is a paradigm shift, treating DR *systemically* in addition to the traditional localized treatments (laser or intravitreal injections).

This study compares the effects of statins and fibrates on diabetic retinopathy (DR), integrating mechanistic, preclinical, and clinical findings. Statins, through HMG-CoA reductase inhibition, reduce LDL cholesterol and exert pleiotropic vascular effects. These include anti-inflammatory, antioxidant, and modest anti-angiogenic actions in retinal tissues. In diabetic models, statins reduce VEGF expression, leukocyte adhesion, and blood-retinal barrier breakdown, suggesting potential for stabilizing early DR.^1,2,6^. However, large trials such as CARDS and HPS found no significant reduction in DR progression or laser treatment rates,^3,7^ likely due to low DR event capture and limited ophthalmic endpoints. Small RCTs have indicated statins may reduce hard exudates or fluorescein leakage,^8,9^ though results were inconsistent.

Fibrates, particularly fenofibrate, activate PPAR-α and have demonstrated potent retinal anti-inflammatory and anti-angiogenic effects. These include suppression of VEGF, ICAM-1, MCP-1, and NF-κB activity, stabilization of the blood-retinal barrier, and reduced leukostasis.^10–12^ In two large randomized trials—FIELD and ACCORD Eye—fenofibrate significantly reduced DR progression and the need for laser photocoagulation, especially among patients with existing DR.^13^ These benefits were independent of LDL levels and were not observed in patients without baseline DR. Observational studies further support fenofibrate’s retinal benefits, including reduced risk of VH, photocoagulation, and anti-VEGF therapy when added to statin treatment.^14,15^

Fibrates have demonstrated greater and more consistent efficacy for DR outcomes than statins. While both drug classes possess vascular protective properties, fibrates act more directly on DR-relevant pathways. Statins may offer microvascular benefit indirectly via systemic vascular improvements, whereas fenofibrate appears to target retinal inflammation and angiogenesis more specifically.^16–18^ In ACCORD Eye, adding fenofibrate to statin therapy reduced the 4-year risk of DR progression by 40%.^3^ This benefit was corroborated in real-world evidence from a Korean cohort of over 70,000 patients with diabetes.^19^

While statins are essential for reducing cardiovascular risk in diabetes, their role in protecting against DR remains less clear. Statins may indirectly benefit retinal health by improving endothelial function and lowering systemic inflammation^20^, but their impact on DR appears modest compared to glycemic and blood pressure control. This may explain why large statin trials did not show significant DR benefits. Retinopathy progression in those studies was largely driven by diabetes duration and metabolic control. It’s also possible that statins require longer use to influence DR, as supported by observational data showing reduced DR incidence in patients on long-term statin therapy.^21^

Another aspect is that statins and fibrates address different metabolic profiles. In type 2 diabetes, mixed dyslipidemia (high triglycerides, low HDL) is common and may be particularly harmful to microvessels. Fenofibrate targets that profile and also reduces diabetic dyslipidemia-related factors like Lp-PLA2 (an enzyme linked to vascular inflammation).^22^ Statins, while excellent for lowering LDL, don’t reduce triglycerides as effectively. This might partially explain why adding fenofibrate yields incremental benefit – it addresses the lipid abnormality (and downstream inflammation) that statins leave behind. From a mechanistic viewpoint, PPAR-α activation by fenofibrate directly modulates genes in retinal cells (for instance, upregulating anti-inflammatory proteins in the endothelium) that statins do not directly influence.^23^ One striking experimental finding is fenofibrate’s ability to inhibit retinal NF-κB and HIF-1, two master regulators of inflammation and angiogenesis respectively. Statins have anti-NF-κB effects too, but fenofibrate’s effect might be stronger or more targeted to the retina’s needs.^24^

In clinical practice, one must also consider safety and tolerability. Both statins and fenofibrate are generally safe, but combination therapy requires monitoring. Fenofibrate can raise serum creatinine and, rarely, can contribute to muscle toxicity especially when combined with a statin (though this was not a major issue in trials).^25^ The decision to add fenofibrate for DR needs to be individualized, balancing the modest risk of side effects against the potential to preserve vision. For a patient with type 2 diabetes and signs of DR (e.g. moderate NPDR) despite good glycemic control, adding fenofibrate could be a prudent step – especially if they also have elevated triglycerides or other indications. For a patient with no retinopathy, fenofibrate might not be necessary purely for DR prevention (as evidence for primary prevention is weaker), unless they need it for lipid control anyway. Statins, of course, should be prescribed per cardiovascular guidelines for cardiovascular prevention in virtually all patients with diabetes; their use is not predicated on DR status, but it’s reassuring that they likely confer some DR benefit on the side.^26^

The advent of intravitreal anti-VEGF injections has revolutionized treatment of diabetic macular edema and proliferative DR. Yet, anti-VEGF therapy can be costly and requires frequent intraocular injections, which carry risks and burdens. If fenofibrate can reduce the need for anti-VEGF by 22% as the Korean study suggests (HR 0.78),^27^ that could translate into fewer injections and less cumulative risk for patients. In this sense, systemic fenofibrate and statins may complement ocular therapies: by reducing the incidence of severe retinal pathology, they may decrease how often patients require laser or injections. This integrated management approach (optimizing systemic factors alongside ocular treatments) may represent future of DR care. The FIELD and ACCORD trials even predated wide anti-VEGF use, so now we have more tools than ever – combining good diabetes care, lipid management, and ocular therapy – to combat DR. Future studies may explore if patients on fenofibrate have better outcomes or need fewer injections in diseases like diabetic macular edema.

One open question is whether other lipid-modifying agents (beyond statins/fibrates) might also benefit DR. For instance, the PCSK9 inhibitors (powerful LDL-lowering antibodies) or omega-3 fatty acids (which can lower triglycerides) might theoretically help. However, no current evidence suggests that PCSK9 inhibitors impact DR (they mainly lower LDL, similar to statins). Omega-3 fatty acids have anti-inflammatory properties, but again no clinical trial data in DR yet. Given fenofibrate’s success, there is interest in newer PPAR-α agonists or dual PPAR agonists that might be even more potent. Also, vitamin B3 (niacin) raises HDL and might have PPAR-α effects; it’s untested in DR but theoretically could be studied. For now, fenofibrate remains the standout agent for retinopathy among lipid drugs.

Ultimately, these findings reinforce the importance of a multifactorial approach to diabetic retinopathy. The Steno-2 trial years ago showed that intensive multifactorial therapy (glucose, BP, lipids) reduced microvascular complications in diabetics. Fenofibrate’s DR benefit is another piece of that puzzle, highlighting lipids as modifiable risk factors for DR progression. Clinicians should ensure that, in addition to controlling blood sugar and blood pressure, any dyslipidemia in a diabetic patient is addressed – not only to prevent heart attacks, but potentially to preserve vision. Given the evidence, if a patient with type 2 diabetes and moderate NPDR is already on a statin (as they should be) and has no contraindications, adding fenofibrate could be considered to slow retinopathy, in consultation with their ophthalmologist. Educating both patients and providers about this indication is important, as awareness is still not universal (fenofibrate is not commonly thought of as an “eye drug,” and many endocrinologists or ophthalmologists may not routinely use it for DR yet). However, as more post-trial data accumulate and as bodies like the ADA possibly integrate this into guidelines, the use of fenofibrate for DR might increase.

## Conclusions

In conclusion, fibrates and statins – originally cardiovascular medications – have shown noteworthy effects on diabetic retinopathy, with fibrates emerging as the more potent intervention for retinal outcomes. Fenofibrate therapy in type 2 diabetes has been proven to slow DR progression and reduce the risk of proliferative disease or the need for retinal laser treatment.^28^ Its mechanisms involve PPAR-α mediated anti-inflammatory and anti-angiogenic actions that directly counteract DR pathogenesis. Statins, while essential for cardiovascular health in diabetes, have a more modest impact on DR: they likely help to a degree (supported by observational data showing lower DR rates in statin users),^29^ but RCT evidence for statins on retinopathy is inconclusive. Combining a statin with fenofibrate yields the lipid benefits of the former and the retinopathy-specific benefits of the latter – an approach validated by trial data.^28^ For patients with type 2 diabetes and established retinopathy, fenofibrate therapy (in addition to standard care) should be considered to reduce the risk of DR progression, as endorsed by various studies and some guidelines.^30^ In contrast, statin therapy should be continued for all its systemic benefits, with the understanding that it may confer some protection but is not a substitute for fenofibrate’s effect on the retina.

Looking forward, the integration of systemic therapies like fenofibrate with routine diabetes care offers a promising avenue to diminish the burden of diabetic retinopathy. By reducing microvascular damage at its source, such treatments can complement ocular therapies and potentially decrease the frequency of invasive procedures like intravitreal injections. The story of statins and fibrates in DR also highlights the complex interplay of metabolic factors in diabetes complications – control of blood sugar, blood pressure, and blood lipids *together* yields the best outcome for retinal health.^31^ Ongoing research may further clarify optimal patient selection (e.g. who benefits most from fenofibrate) and whether newer agents can replicate or surpass these effects. For now, the available evidence solidly supports fenofibrate as an efficacious option to slow diabetic retinopathy progression, marking a significant achievement in the medical management of this blinding disease. With comprehensive care that includes attention to dyslipidemia, we stand a better chance of preserving vision in our patients with diabetes.

## Data Availability

The data that support the findings of this study are derived from the TriNetX Global Collaborative Network, which contains de-identified patient information from participating healthcare organizations. Due to data use agreements, the raw data are not publicly available. Access to the TriNetX platform and its datasets can be obtained by qualified researchers through a request to TriNetX (https://www.trinetx.com), subject to institutional approval and data sharing policies.

## Author Contributions

*Concept and design:* Luke Nelson, Jesse Maynard, Matthew Morckos, Miriam Michael,MD, Salman Yousuf, DO

*Acquisition, analysis, or interpretation of data:* Luke Nelson, Jesse Maynard, Matthew Morckos, Miriam Michael,MD, Salman Yousuf, DO

*Drafting of the manuscript:* Luke Nelson, Jesse Maynard, Matthew Morckos, Ruben Petit Homme, PhD, Samrawit Zinabu, MD, Miriam Michael, MD, Salman Yousuf, DO

*Critical review of the manuscript for important intellectual content:* Luke Nelson, Jesse Maynard, Matthew Morckos, Ruben Petit Homme, PhD, Samrawit Zinabu, MD, Miriam Michael, MD, Salman Yousuf, DO

*Statistical analysis:* Luke Nelson, Jesse Maynard, Samrawit Zinabu, MD, Rawan Elkomi

*Administrative, technical, or material support:* Rawan Elkomi, Samrawit Zinabu, MD, Miriam Michael, MD, Salman Yousuf, DO

*Supervision:* Miriam Michael, MD, Salman Yousuf, DO

## Funding/Support

No funding source was used for this research. The authors received no financial support for conducting this study, analyzing the data, or preparing this manuscript. Beyond usual salary, no one received financial compensation for their contribution.

## Disclosures

The authors have no relevant financial or non-financial relationships to disclose.

## Non-standard Abbreviations and Acronyms

DR: Diabetic retinopathy
T2DM: Type 2 Diabetes Mellitus
NPDR: nonproliferative diabetic retinopathy
VH: vitreous hemorrhage
NV: neovascularization
TRD: tractional retinal detachment
NVG: neovascular glaucoma
PPV: pars plana vitrectomy
HMG-CoA: 3-Hydroxy-3-Methylglutaryl-Coenzyme A
PPAR-α: Peroxisome proliferator-activated receptor alpha
HCOs: healthcare organizations
wet AMD: exudative age-related macular degeneration
LLD: lipid-lower drug
PRP: panretinal, photocoagulation
PDR: proliferative diabetic retinopathy
AST: aspartate aminotransferase
ALT: alanine aminotransferase
AKI: acute kidney injury
RR: risk ratio
CI: confidence intervals
HR: Hazard ratios

## Notes

### Competing Interest Statement

The authors have declared no competing interest.

### Funding Statement

The authors received no financial support for the research, authorship, or publication of this article. No external funding, grants, or in-kind support were obtained for this study.

### Author Declarations

This study was conducted using deidentified patient data from the TriNetX Global Collaborative Network. Per the Health Insurance Portability and Accountability Act (HIPAA) Privacy Rule {section sign}164.514(b)(1), a qualified expert determined that the use of these data is exempt from institutional review board (IRB) oversight. No patient identifiers were accessed, and all analyses complied with relevant ethical guidelines.

## References

1. Ioannidou E, Tseriotis V-S, Tziomalos K. Role of lipid-lowering agents in the management of diabetic retinopathy. World J. Diabetes. 2017;8:1–6.

2. Karti O, Saatci AO. Fenofibrate and diabetic retinopathy. Med. Hypothesis Discov. Innov. Ophthalmol. 2024;13:35–43.

3. Wright AD, Dodson PM. Medical management of diabetic retinopathy: fenofibrate and ACCORD Eye studies. Eye Lond. Engl. 2011;25:843–849.

4. Noonan JE, Jenkins AJ, Ma J-X, Keech AC, Wang JJ, Lamoureux EL. An update on the molecular actions of fenofibrate and its clinical effects on diabetic retinopathy and other microvascular end points in patients with diabetes. Diabetes. 2013;62:3968–3975.

5. Diabetic retinopathy: management and monitoring [Internet]. London: National Institute for Health and Care Excellence (NICE); 2024 [cited 2025 May 9]. Available from: http://www.ncbi.nlm.nih.gov/books/NBK607261/

6. Gupta A, Gupta V, Thapar S, Bhansali A. Lipid-lowering drug atorvastatin as an adjunct in the management of diabetic macular edema. Am. J. Ophthalmol. 2004;137:675–682.

7. Sen K, Misra A, Kumar A, Pandey RM. Simvastatin retards progression of retinopathy in diabetic patients with hypercholesterolemia. Diabetes Res. Clin. Pract. 2002;56:1–11.

8. Kawasaki R, Konta T, Nishida K. Lipid-lowering medication is associated with decreased risk of diabetic retinopathy and the need for treatment in patients with type 2 diabetes: A real-world observational analysis of a health claims database. Diabetes Obes. Metab. 2018;20:2351–2360.

9. Shiono A, Sasaki H, Sekine R, Abe Y, Matsumura Y, Inagaki T, Tanaka T, Kodama T, Aburatani H, Sakai J, et al. PPARα activation directly upregulates thrombomodulin in the diabetic retina. Sci. Rep. 2020;10:10837.

10. Chen Y, Hu Y, Lin M, Jenkins AJ, Keech AC, Mott R, Lyons TJ, Ma J. Therapeutic Effects of PPARα Agonists on Diabetic Retinopathy in Type 1 Diabetes Models. Diabetes. 2013;62:261–272.

11. Mozetic V, Pacheco RL, Latorraca C de OC, Riera R. Statins and/or fibrates for diabetic retinopathy: a systematic review and meta-analysis. Diabetol. Metab. Syndr. 2019;11:92.

12. Vail D, Callaway NF, Ludwig CA, Saroj N, Moshfeghi DM. Lipid-Lowering Medications Are Associated with Lower Risk of Retinopathy and Ophthalmic Interventions among United States Patients with Diabetes. Am. J. Ophthalmol. 2019;207:378–384.

13. Fenofibrate therapy in reducing the progression of diabetic retinopathy: revisiting the FIELD and ACCORD-EYE studies through the LENS trial | Eye [Internet]. [cited 2025 May 9];Available from: https://www.nature.com/articles/s41433-024-03410-9

14. Meer E, Bavinger JC, Yu Y, Hua P, McGeehan B, VanderBeek BL. Statin Use and the Risk of Progression to Vision Threatening Diabetic Retinopathy. Pharmacoepidemiol. Drug Saf. 2022;31:652–660.

15. Kataoka SY, Lois N, Kawano S, Kataoka Y, Inoue K, Watanabe N. Fenofibrate for diabetic retinopathy. Cochrane Database Syst. Rev. 2023;>2023:CD013318.

16. Diabetic Retinopathy Tx: A Role for Fibrates and Statins? [Internet]. Am. Acad. Ophthalmol. 2019 [cited 2025 May 9];Available from: https://www.aao.org/eyenet/article/diabetic-retinopathy-tx-fibrates-and-statins

17. Fiévet C, Staels B. Combination therapy of statins and fibrates in the management of cardiovascular risk. Curr. Opin. Lipidol. 2009;20:505–511.

18. Jacobson TA, Zimmerman FH. Fibrates in Combination With Statins in the Management of Dyslipidemia. J. Clin. Hypertens. 2007;8:35–41.

19. Lim J, Choi Y-J, Kim BS, Rhee T-M, Lee H-J, Han K-D, Park J-B, Na JO, Kim Y-J, Lee H, et al. Comparative cardiovascular outcomes in type 2 diabetes patients taking dapagliflozin versus empagliflozin: a nationwide population-based cohort study. Cardiovasc. Diabetol. 2023;22:188.

20. Kawasaki R, Konta T, Nishida K. Lipid-lowering medication is associated with decreased risk of diabetic retinopathy and the need for treatment in patients with type 2 diabetes: A real-world observational analysis of a health claims database. Diabetes Obes. Metab. 2018;20:2351–2360.

21. Role of lipid-lowering agents in the management of diabetic retinopathy - PMC [Internet]. [cited 2025 May 9];Available from: https://pmc.ncbi.nlm.nih.gov/articles/PMC5237812/

22. Fenofibrate and diabetic retinopathy - PMC [Internet]. [cited 2025 May 9];Available from: https://pmc.ncbi.nlm.nih.gov/articles/PMC11227662/

23. Abcouwer SF. Direct Effects of PPARα Agonists on Retinal Inflammation and Angiogenesis May Explain How Fenofibrate Lowers Risk of Severe Proliferative Diabetic Retinopathy. Diabetes. 2013;62:36–38.

24. Garcia-Ramírez M, Hernández C, Palomer X, Vázquez-Carrera M, Simó R. Fenofibrate prevents the disruption of the outer blood retinal barrier through downregulation of NF-κB activity. Acta Diabetol. 2016;53:109–118.

25. Efficacy and Safety of Fenofibrate-Statin Combination Therapy in Patients With Inadequately Controlled Triglyceride Levels Despite Previous Statin Monotherapy: A Multicenter, Randomized, Double-blind, Phase IV Study - ScienceDirect [Internet]. [cited 2025 May 9];Available from: https://www.sciencedirect.com/science/article/pii/S0149291821003027

26. Eldor R, Raz I. American Diabetes Association Indications for Statins in Diabetes. Diabetes Care. 2009;32:S384–S391.

27. Another Study Shows Fenofibrate Benefit on Diabetic Retinopathy [Internet]. Medscape. [cited 2025 May 9];Available from: https://www.medscape.com/viewarticle/981317

28. Medical management of diabetic retinopathy: fenofibrate and ACCORD Eye studies - PubMed [Internet]. [cited 2025 May 9];Available from: https://pubmed.ncbi.nlm.nih.gov/21436845/

29. Lipid-Lowering Medications Are Associated with Lower Risk of Retinopathy and Ophthalmic Interventions among United States Patients with Diabetes - PubMed [Internet]. [cited 2025 May 9];Available from: https://pubmed.ncbi.nlm.nih.gov/31194953/

30. Practitioners TRAC of general. The use of fenofibrate in the management of patients with diabetic retinopathy: an evidence-based review [Internet]. Aust. Fam. Physician. [cited 2025 May 9];Available from: https://www.racgp.org.au/afp/2015/june/the-use-of-fenofibrate-in-the-management-of-pa-2

31. Ioannidou E, Tseriotis V-S, Tziomalos K. Role of lipid-lowering agents in the management of diabetic retinopathy. World J. Diabetes. 2017;8:1–6.

